# Estimation of Undetected Symptomatic and Asymptomatic cases of COVID-19 Infection and prediction of its spread in USA

**DOI:** 10.1101/2020.06.21.20136580

**Authors:** Ashutosh Mahajan, Ravi Solanki, Namitha Sivadas

## Abstract

The reported COVID-19 cases in the USA have crossed over 2 million, and a large number of infected cases are undetected whose estimation can be done if country-wide antibody testing is performed. In this work, we estimate this undetected fraction of the population by modeling and simulation approach. We propose a new epidemic model SIPHERD in which three categories of infection carriers Symptomatic, Purely Asymptomatic, and Exposed are considered with different transmission rates that are taken dependent on the lockdown conditions, and the detection rate of the infected carriers is taken dependent on the tests done per day. The model is first validated for Germany and South Korea and then applied for prediction of total number of confirmed, active and death, and daily new positive cases in the United States. Our study also demonstrates the possibility of a second wave of the infection if social distancing regulations are relaxed to a large extent. We estimate that around 12.7 million people are already infected, and in the absence of any vaccine, 17.7 million (range: 16.3-19.2) people, or 5.3% (range: 4.9–5.8) of the population will be infected by when the disease spread ends in the USA. We find the Infection to Fatality Ratio to be 0.93% (range: 0.85-1.01).

## I. Introduction

The outbreak of pandemic Coronavirus disease 2019 (COVID-19) has led to more than 8 million total reported infections and 450 thousand deaths worldwide [1], and serious efforts are needed for its containment. The Coronavirus SARS-CoV-2 has affected not just the public health but made a drastic impact on the economy of the world as well, due to the lockdown situations in many countries, including the United States of America. In the USA, the first positive case of COVID-19 is reported on January 20, 2020, in a man who returned from Wuhan, China, where the outbreak was first identified, and the first death took place in the first week of February [2]. A major control measure was announced on March 16, restricting the gatherings of more than ten people. However, the COVID 19 spread to almost 50 states throughout the country by March-end [1], [3]. Now, USA has become the most affected country in terms of confirmed, active, and death cases in the world [1].

Pandemics have hit humanity many times in the past also, and mathematical models are already available for infectious diseases [4], [5], [6], [7]. Mathematical modeling of the epidemic has an unavoidable role in helping the healthcare sector by predicting the hospital requirements in advance and for setting up the critical care systems for the patients [8] [9].Modeling and simulation can also predict the extent of the contagious disease and helps the administration of the nations in decision making to implement radical control measures for its containment [10]. In order to devise the lockdown strategy, it is imperative that the prediction of the disease spread is available to the policymakers.

COVID-19 is different from the previously known SARS (Severe acute respiratory syndrome) infection, with features such as the existence of purely asymptomatic cases [11] and the spread of the infection from those as well as from the exposed ones in the incubation period [12]. Our proposed mathematical model incorporates the above facts for the COVID-19 epidemic. Many other epidemiological models are proposed for the COVID-19 infection spread [13], [14], [15], [16], [17], [18].

In [18], Murray and his collaborators predicted the number of hospital beds that will be needed, critical health care requirements like ICU and ventilators based on the data of present COVID-19 patients and the total number of deaths in the United States and the European Economic Area.

In this paper, we formulate a mathematical model, named SIPHERD for the COVID-19 epidemic and apply it for forecasting the number of total active and confirmed cases, daily new positive and death cases in the USA, according to the conditions of the lockdown and the number of tests performed per day.

## II. Methods

### A. Mathematical Model SIPHERD

We model the dynamics of the COVID-19 disease spread by dividing the population into different categories, as listed below.

- S - fraction of the total population that is healthy and has never caught the infection
- E - fraction of the total population that is exposed to infection, transmit the infection and turn into either Symptomatic or purely Asymptomatic, and not detected
- I - fraction of the total population infected by the virus that shows symptoms and undetected
- P - fraction of the total population infected by the virus that doesn’t show symptoms even after the incubation period and undetected. These are the purely Asymptomatic cases
- H - fraction of the total population that are found positive in the test and either hospitalized or quarantined
- R - fraction of the total population that has recovered from the infection
- D - fraction of the total population that are deceased due to the infection.

We model the dynamics of the COVID-19 disease spread by dividing the population into different categories, as Susceptible (S), Exposed (E), Symptomatic (I), Purely Asymptomatic (P), Hospitalized or Quarantined (H), Recovered (R) and Deceased (D). The SIPHERD model equations are a set of coupled ordinary differential equations (1 to 7) for the defined entities (S,I,P,H,E,R,D). As seen in Fig.1, the rates of transfer from one category to another are the model parameters, and a set of differential equations for the entity in each category is formed.

**Fig. 1:**
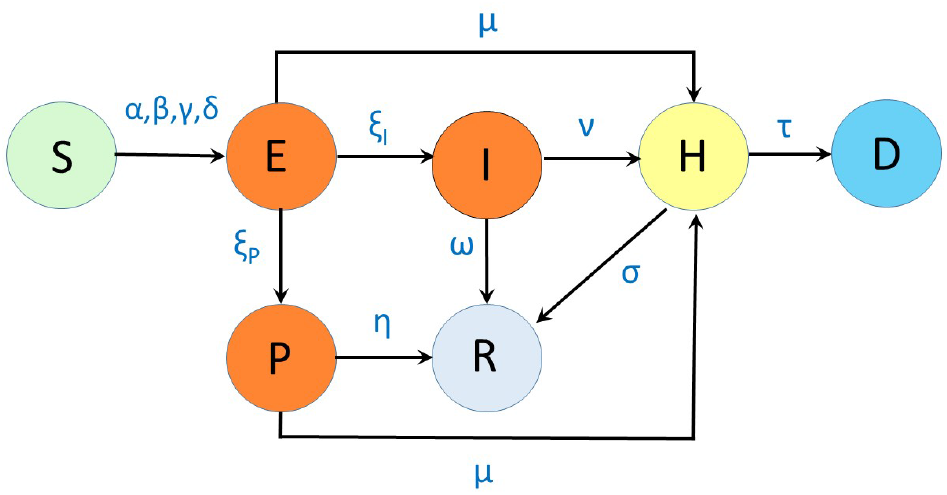
Schematic of the SIPHERD Model: *α, β, γ, δ* are rates of transmission of infection; *ξ*_*I*_, *ξ*_*P*_ are rates of transfer from exposed to Symptomatic and Asymptomatic; *ω, η, s* are recovery rates; *µ, ν* are detection rates, and *τ* is mortality rate.

The probability of getting the infection is assumed uniform among the susceptible people, although the disease spreads localized in hot-spots. Therefore, even though the disease has spread very differently in different US states, the model considers ‘average effect’ for the estimation of the infected and death cases.

We write the model equations that are independent of the population of the country by considering the fraction of the people in each category[19]. The various rates listed in TABLE I are the parameters of the problem which are not known, and only possible range is available and the initial conditions E(0), P(0) and I(0) are also not exactly known. Some of the parameters such as rates of infection (*α, β, γ*) change with time in steps, depending on the lockdown and social distancing conditions, and probability rate of detection (*ν*) changes with time depending on tests per day (*T*_*PD*_). The model equations are written as,

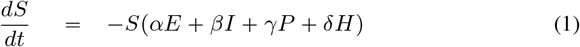

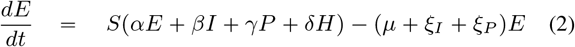

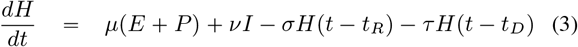

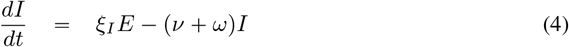

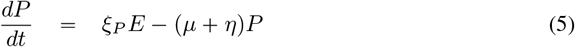

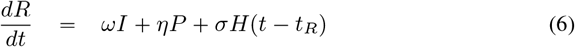

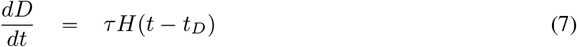

where, *t*_*R*_ and *t*_*D*_ are the delay associated with the recovery and death respectively with respect to active cases *H*. We have taken into account this delay because the active cases are reported after the testing and admission to healthcare or quarantine center, and the number of recovery and death of the admitted will not immediately follow the active or *H* category number. All fractions add up to unity that can also be seen from summing the above equations.

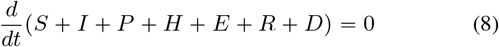

**TABLE I.**
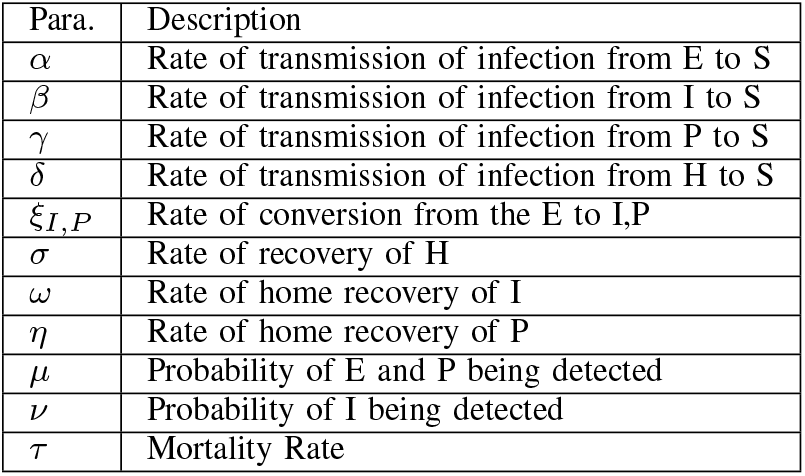

The basic reproduction number (*R*_0_) can be written by observing the inflow and outflow rates for each infectious category (E,I,P,H) shown in Fig 1 of the manuscript. The contribution of each of these categories for the reproduction number can be written as the ratio of the sum of inflow rates and the sum of outflow rates multiplied by the rate of transmission of infection of that category.

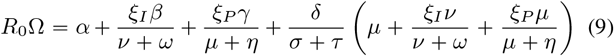

where, Ω = *µ* + *ξ*_*I*_ + *ξ*_*P*_.

As the existence of Purely Asymptomatic cases is a distinct feature of COVID-19, and it is crucial to identify the proportion of such cases among the total infected in order to build a realistic model. The *Diamond Princess* Cruise study is the key to identify the proportion of Asymptomatic cases as all the susceptible people onboard were tested. The asymptomatic proportion of the infected persons onboard the *Diamond Princess* Cruise is estimated in [11]. Among the 634 tested positive onboard, 328 were found asymptomatic, i.e. more than 50 percent of the confirmed cases were not showing any specific symptoms of COVID-19. The ratio of purely Asymptomatic (P) to total Asymptomatic (E+P) cases is reported to be 0.35, and the ratio of purely Asymptomatic to the total infected (E+P+I) is 0.179 [11].

The above-observed ratios can be written in terms of the entities on the Cruise (with a bar) as all the people onboard were tested,

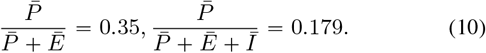

This implies that 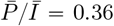. These reported numbers are used to fix the proportion between *ξ*_*P*_ and *ξ*_*I*_ as 0.36 and the proportion of initial conditions *E*(0), *I*(0) and *P*(0) as well. In other words, out of 136 exposed cases, after the incubation, 36 will turn to be purely asymptomatic, and 100 will have symptoms.

The detection of the Asymptomatic and Symptomatic cases can be taken dependent on the number of tests done per day (*T*_*PD*_). For the Symptomatic cases, the detection is more probable as the infected person can approach for the tests and more likely to be tested. The detection of Symptomatic is taken in two parts, a constant (*ν*_0_) and another part proportional to the tests done per day. This can be written in terms of parameters as,

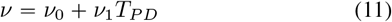

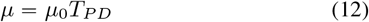

where, *µ*_0_, *ν*_0_, and *ν*_1_ are positive constants. The total confirmed cases are the addition of the active cases, extinct cases, and a part of the recovered that were detected. This can be written as

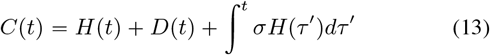

Asymptomatic carriers of Coronavirus-nCoV2 do transmit the disease. Also, the infection can be transmitted from the person who is not showing illness during the incubation period [12]. This can be included in the model by considering *E* category people and their transmission rate *α*. Hospitalized and quarantined cases can also transmit the disease, and this low rate is taken as parameter *δ*. We model the transmission rate of infection change with time in steps, depending on the conditions of the lockdown. The detection of the Asymptomatic and Symptomatic cases is taken dependent on the number of tests done per day (*T*_*PD*_). For the Symptomatic cases, the detection is more probable as the infected person can approach for the tests and more likely to be tested. The transfer rate from E to I (*ξ*_*I*_) is the inverse of the incubation period, whose mean is reported 5.2 days [20]. Recovery time of Symptomatic cases is taken as 14 days. The total confirmed cases are the addition of the active cases, extinct cases, and a part of the recovered that were detected. The rate of transmission of infection from the Asymptomatic carrier (*α, γ*) for a country is typically taken higher than the Symptomatic ones (*β*) as the Asymptomatic carrier may not be aware of his/her infection, and Susceptible may not be keeping distance as no symptoms are seen. The mortality rate (*τ*) is taken differently for different countries as it depends on the immunity and how effectively the critical patients are taken care of by the hospitals.

### B. Optimization of the Parameters

Some of the parameters namely, *ω, η, ξ*_*P*_, *ξ*_*I*_ have fixed value as those represent the characteristics of the disease itself. The remaining parameters are to be obtained that generate the evolution of the dynamical system close to the actual data. Manual tuning of the parameters for the best fit is quite a tedious task. For this purpose, we write a cost function in terms of the standard deviation from the actual data and model data for the confirmed and the active cases as the following

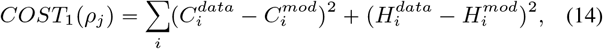

where, *ρ*_*j*_ are the different parameters that are to be obtained. For estimation of the undetected infected cases for the USA, we write another cost for the first 40 days from March 1, 2020

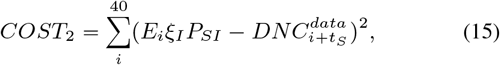

where, *P*_*SI*_ is the probability of the daily new symptomatic cases develop severe symptoms after time a delay of *t*_*S*_ days and were reported as daily new cases (DNC). The net cost function is sum of *COST*_1_ and *COST*_2_.

A MATLAB function *‘fmincon’* is used to find the minimum of a problem depending on a set of parameters that can have upper and lower bounds. *fmincon* returns the set of parameters within the given range, which minimizes the COST function defined above. As there could be multiple sets of parameters giving out ‘good fit’ to the real data, other physical constraints on the parameter sets can be considered. One of them is a reasonable value of the reproduction number. Secondly, the rate of transmission of infection before lockdown has to be greater than after lockdown. The mortality rate (*τ*) is not optimized but rather calculated directly by the daily number of deaths data (DND) and the active data. The mortality rate for a particular day can be obtained as follows.

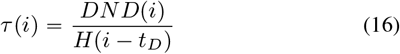

### C. Numerical Implementation and Simulation

The set of coupled equations for the model for a given set of parameters and initial values is solved numerically by dde23 solver routine of MATLAB for ordinary differential equations with a time lag in functions. The non-trivial part is the accurate determination of the parameters that will mimic the situation on the ground. The mathematical problem is to take into account the four actual data sets of the total number of confirmed cases, active cases on a particular day, cumulative deaths and tests done per day and find the set of parameters that will provide the best possible match between the data and model. The extraction of the parameters is automated so that the model can be run on data for various countries. The minimizer of the cost is found to obtain the optimized set of parameters that best fit with the data available till date. The model and the optimization scheme is implemented in MATLAB.

The parameters determined by our model are listed in TABLE II for the countries we studied.

**TABLE II:** Parameters values for the Countries studied

| Para | Germany | S. Korea | USA |
| --- | --- | --- | --- |
| Popul. N | 8.30E7 | 5.10E7 | 3.31e8 |
| $T_0$ | Feb 21 | Feb 15 | Mar 1 |
| $T_{LD}$ | 29,37 | 18 | 25,40 |
| $\alpha$ (bf. LD) | 0.35 | 0.45 | 0.37 |
| $\alpha$ (af. LD) | 0.23,0.14 | 0.17 | 0.05,0.05 |
| $\beta$ (bf. LD) | 0.19 | 0.16 | 0.31 |
| $\beta$ (af. LD) | 0.18,0.11 | 0.11 | 0.051,0.064 |
| $\gamma$ (bf. LD) | 0.35 | 0.45 | 0.37 |
| $\gamma$ (af. LD) | 0.23,0.14 | 0.17 | 0.05,0.05 |
| $\delta$ | 9E-3 | 5.9E-3 | 6E-3 |
| $\xi_I$ | 0.2 | 0.2 | 0.2 |
| $\xi_P$ | 0.072 | 0.072 | 0.072 |
| $\mu_0$ | 1.67E-6 | 2.05E-5 | 0 |
| $\nu_0$ | 0.05 | 0.07 | 0.023 |
| $\nu_1$ | 4.7E-6 | 9.7E-5 | 5.5E-9 |
| $\omega$ | 0.07 | 0.07 | 0.07 |
| $\eta$ | 0.1 | 0.1 | 0.1 |
| $\sigma$ | 0.065 | 0.034 | 0.015 |
| $t_R$ | 6 | 11 | 10 |
| $t_D$ | 6 | 1 | 1 |
| Ini. Inf. | 100 | 150 | 1000 |

For USA, the rate of transmission of infection is taken to change in three steps. This is done by plotting the total number of cases in log scale and seeing the changes in the slopes and correlating with the government’s regulations on social activities. As seen in Appendix Fig.7 b, we fit the actual mortality rate in steps. It can be seen that the mortality rate improved with time from 2.6% to 0.7%. The mortality rate is expected to improve further, as mild cases will also be reported with more tests available. We estimate the improvement in the mortality rate by calculating the fraction of mild and severe cases and assuming that all mild cases are going to recover. The probability of Symptomatic patients developing severe symptoms (*P*_*SI*_) such as breathlessness, high fever etc has definitely approached the test and was tested in the initial phases. Data from cases reported from 49 states, the District of Columbia, and three U.S. territories (5) to CDC from February 12–March 16 shows that 20.7 reported cases were hospitalized [21]. COVID-NET regions show it to be 21.4 % till April 4 [22], [23] and IHME data March 5-April 4 shows that to be 20.3 % [24] [18]. The estimation of the total infected to hospitalized is reported to be 3.6% in another study for France [25]. Therefore, we estimate the total 17% (3.6/.21) of the total infected develop symptoms that are not mild and are tested and reported in the initial days. In China, this number of non-mild cases is reported to be 19% [26]. For the first 40 days, we put a COST for the above condition that every day, 20% (*ξ*_*I*_) of the exposed develop symptoms, and 17% of them are reported as daily new cases with an average delay of five days. It can be seen from Appendix Fig.8 d that this condition is indeed satisfied as the model curve and real data overlap for the first few days. Later, the gap between the two curves widens as more tests were made available and mild cases also tested.

The projection for the total infected persons is strongly dependent on the value of *P*_*SI*_, which we estimate to be 17% as discussed above. We simulate two more situations for the *P*_*SI*_ value 15% and 19% and plot the time dependence of the Susceptible and Extinct cases in Appendix Fig.8.

### D. Data Collection

We collected the data from the following publicly available data sources: The total number of cases, active cases, daily new cases, and total and daily new deaths is collected from the worldometer [1]. Test per day data is collected from [27].Hospitalization data is collected from [21] [22] [24].

The day on which lockdown is imposed in a country is also taken into account as changes in the slopes of the data for confirmed cases are observed according to it.

## III. Results and Discussion

We apply the SIPHERD model to South Korea and Germany for testing the predictive capability of our model as the disease has almost reached the end stages in these countries. We used the data only for the first 20 and 40 days, respectively, i.e., till March 5 and March 31, and compared the future evolution generated by the model with the actual data, as shown in the grey region in Fig. 2a for South Korea and in Fig.2b for Germany. Parameters extracted by the model from the actual data for the countries studied are listed in TABLE II.

**Fig. 2:**
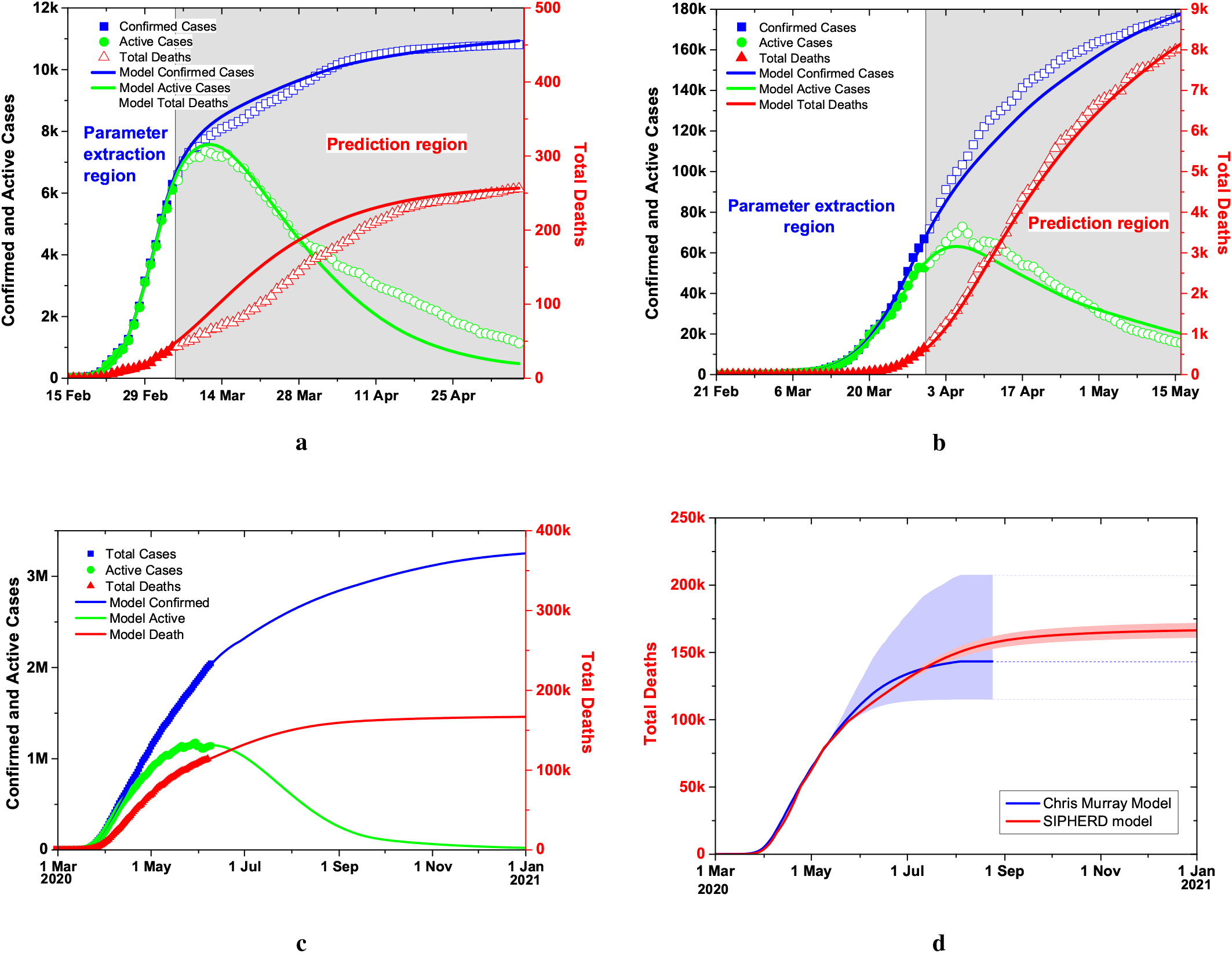
Model predictions for South Korea, Germany and USA. **a**. South Korea data given to the simulator up-to March 5, and thereafter, model comparison with the actual data for the confirmed, active cases and total deaths.**b**. Model prediction using the Germany data up-to March 31 and comparison with the actual data for the confirmed, active cases and total deaths. **c**. Comparison of the Model prediction for USA if the social distancing and lockdown conditions kept same and relaxed after June 15 while the tests increased by 10k daily for both scenarios. **d**. SIPHERD Model compared with ‘Cris Murray Model’ for the projection of the extinct cases.

**Fig. 3:**
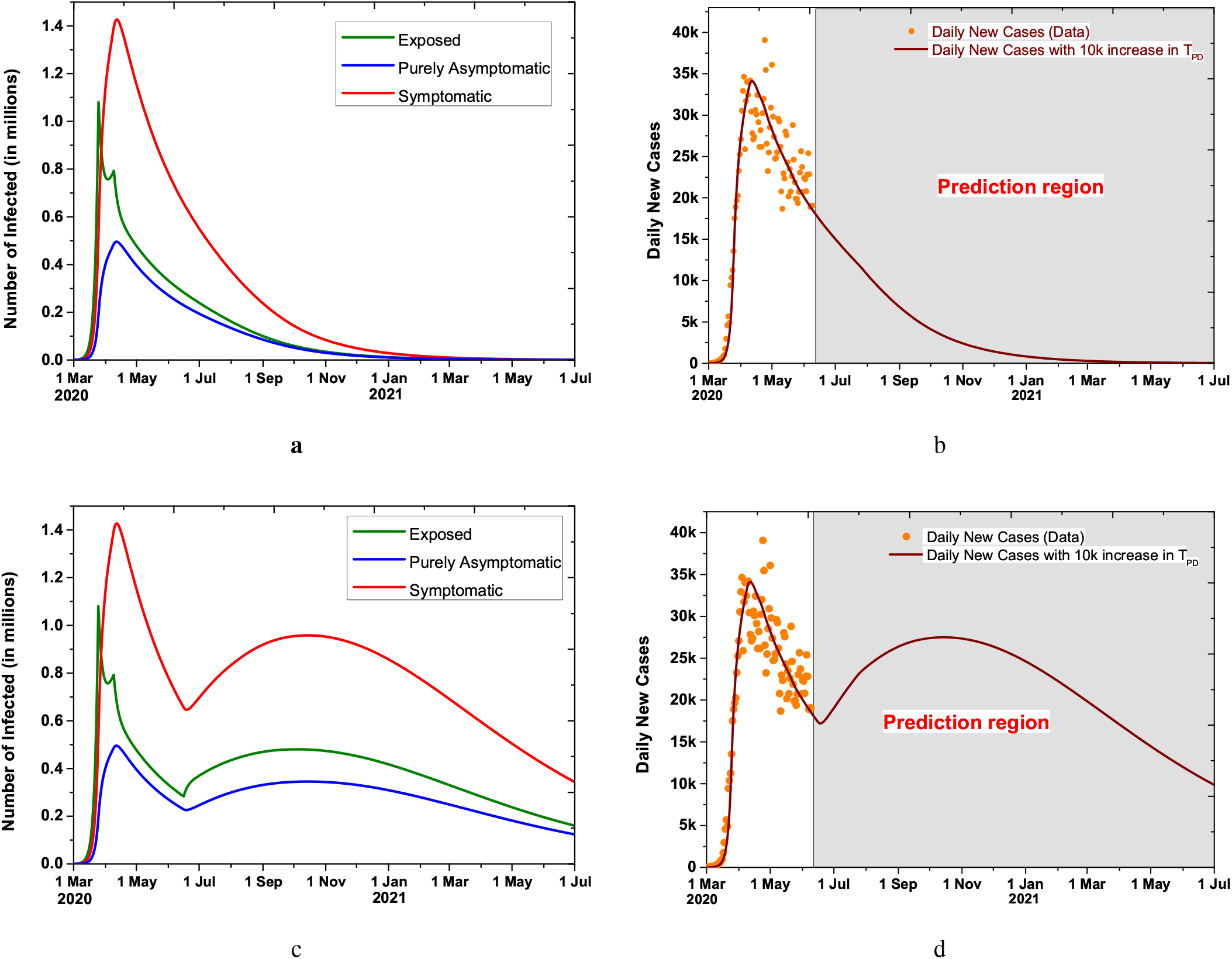
Possibility of a Second Wave of Infection. **a**. The evolution of the undetected number of Infected in Exposed (E), Symptomatic (I), and purely Asymptomatic (P) category with lockdown conditions kept the same. **b**. Model prediction for daily new cases for USA with the increase of 10k tests per day with lockdown conditions kept the same. **c, d**. The evolution of the undetected number of Infected in Exposed (E), Symptomatic (I) and purely Asymptomatic (P) category and Daily new cases when lockdown situations relaxed after June 15 to the extent that rate of transmission of infection increases by 30%.

### A. Effect of Lockdown

The model, thus validated, is then applied to the existing data of the USA for the prediction of the next 500 days, i.e., till July 2021, as shown in Fig.2c. Two scenarios are considered for lockdown and social distancing conditions. One possible scenario is that the conditions are kept the same, and the second one is that they are relaxed after June 15. Test per day assumed to be increased by 10k, which is close to the current trend and taken saturated at 1 million for both the scenarios. The increase in the transmission rate if the lockdown is relaxed is taken as 20% from the current value. The recovery rate *σ* is taken improved after May 26, and the mortality rate is calculated from the data and is improved in steps from initial value 2.65% on March 1 to 0.78% on June 9 as seen in Appendix Fig.7.b. A comparison of the two scenarios is plotted in Fig. 4b.

**Fig. 4:**
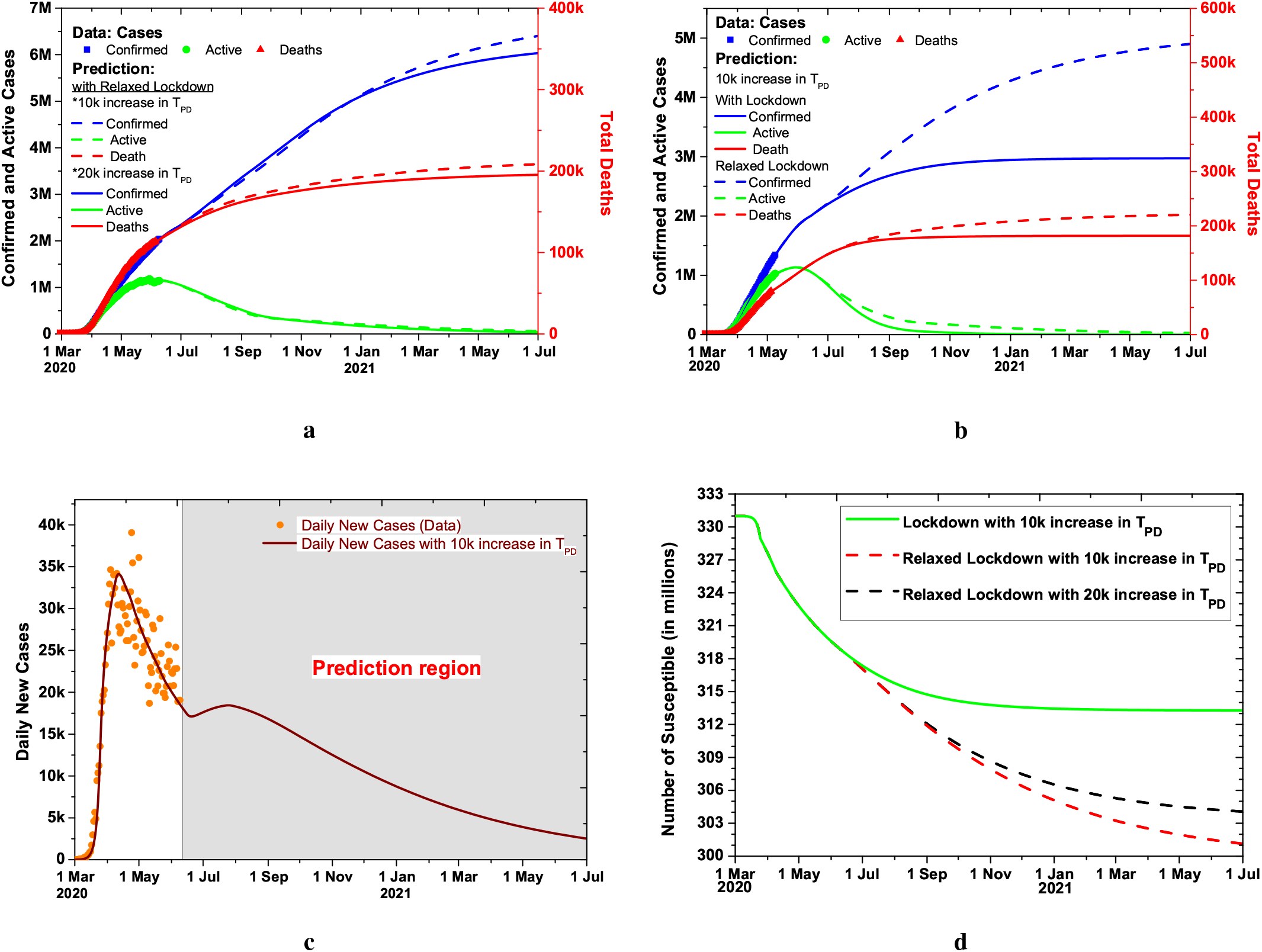
Effect of increased tests and relaxed lockdown. **a**. Effect of increased testing on the total, active and extinct cases, comparison of 10k and 20k increase in Tests per day (*T*_*PD*_) saturated at 1 million and 2 million respectively **b**. Comparison of the Total, Active and extinct cases in lockdown and relaxed lockdown situation **c**. Model prediction for daily new cases with the increase of 10k tests per day with lockdown conditions relaxed after June 15 **d**. The evolution of the Susceptible when lockdown situations kept same and relaxed after June 15 with tests per day increase by 10k and 20k.

The projection of the “Chris Murray” model is compared with SIPHERD model for the total number of deaths in Fig.2d. The prediction range of “Chris Murray” Model can be seen large compared to SIPHERD model.

### B. Estimation of the undetected Cases

Since only symptomatic cases were tested, the detection probability of Asymptomatic (*µ*) is taken zero. There can be many parameter sets, including the probability rate of detection of Symptomatic *ν*, that give a good match between the simulation results and the actual data. It is, therefore, not possible to know the exact number of undetected infected people merely by fitting the model curves with the data. The value of *ν* is fixed by the characteristics of the disease, which is the ratio of severe and mild cases. Most of the COVID-19 patients show mild symptoms, and the number of symptomatic patients that are considered severe is taken 17% of the total Symptomatic. In the initial days of the spread of the infection, due to the lack of test availability, only severe cases were tested. This fact can be used to get an estimate of the undetected symptomatic cases. For illustration, out of 1000 exposed (E) cases, 20% (200) reach Symptomatic (I) category in a day as *ξ*_*I*_ 0.2, and out of those after an average delay of five days, 17% (34) become severe cases and reported as daily new cases in the initial days. This relationship between the available real data of Daily new cases and exposed category number in the initial days gives a constraint on the estimated Exposed cases.

Application of this constraint in the model equations shows that the peak number of undetected Symptomatic infected people go upto 1.4 million. The time evolution of the totally unknown and undetected part of the infected categories for USA is plotted in Fig. 3a. As shown in Fig. 4d, the total number of Susceptible can be around 313 million by the end of this year, assuming that the conditions on lockdown and social distancing remain the same and vaccine is not introduced. Till June 15, 12.61 million, i.e. 3.81% of the USA population could be infected, and this number will increase to 17.71 million people, which is 5.35% of the total population when the infection ends according to the model projections. This number is surprisingly close to the estimated number by large scale antibody testing in Spain [28].

### C. Second Wave Possibility

If the social distancing norms are relaxed from June 15, 2020, to the extent that the current rate of transmission of infection increase by 30%, then our model simulation shows that there is a possibility of the second wave of infection spread as seen in Fig.3c,d. The number of tests is taken increasing 10k every day for this study.

Our study finds that the infection to fatality ratio for USA could be around 0.93 (see Appendix Fig.8 c), which is slightly higher than reported in [25]. This could be due to a lower estimate of the improvement in mortality rate in the coming months. The initial reproduction number 4.8 on March 1 came down to a value of 1.2 on April 1, and to 0.8 on April 15, as seen in Appendix Fig.7 f.

The recovery rate of the Active (*H*) category for USA is found to be low compared to South Korea or Germany, which may be attributed to either incorrect reporting of the Active cases [29] or the testing of serious cases only and lesser recovery in hospitals compared to quarantined with mild symptoms. It can also be seen in Appendix Fig.5 d and 6d that compared to South Korea and Germany, the Symptomatic cases dominate the infected in USA rather than the Exposed.

**Fig. 5:**
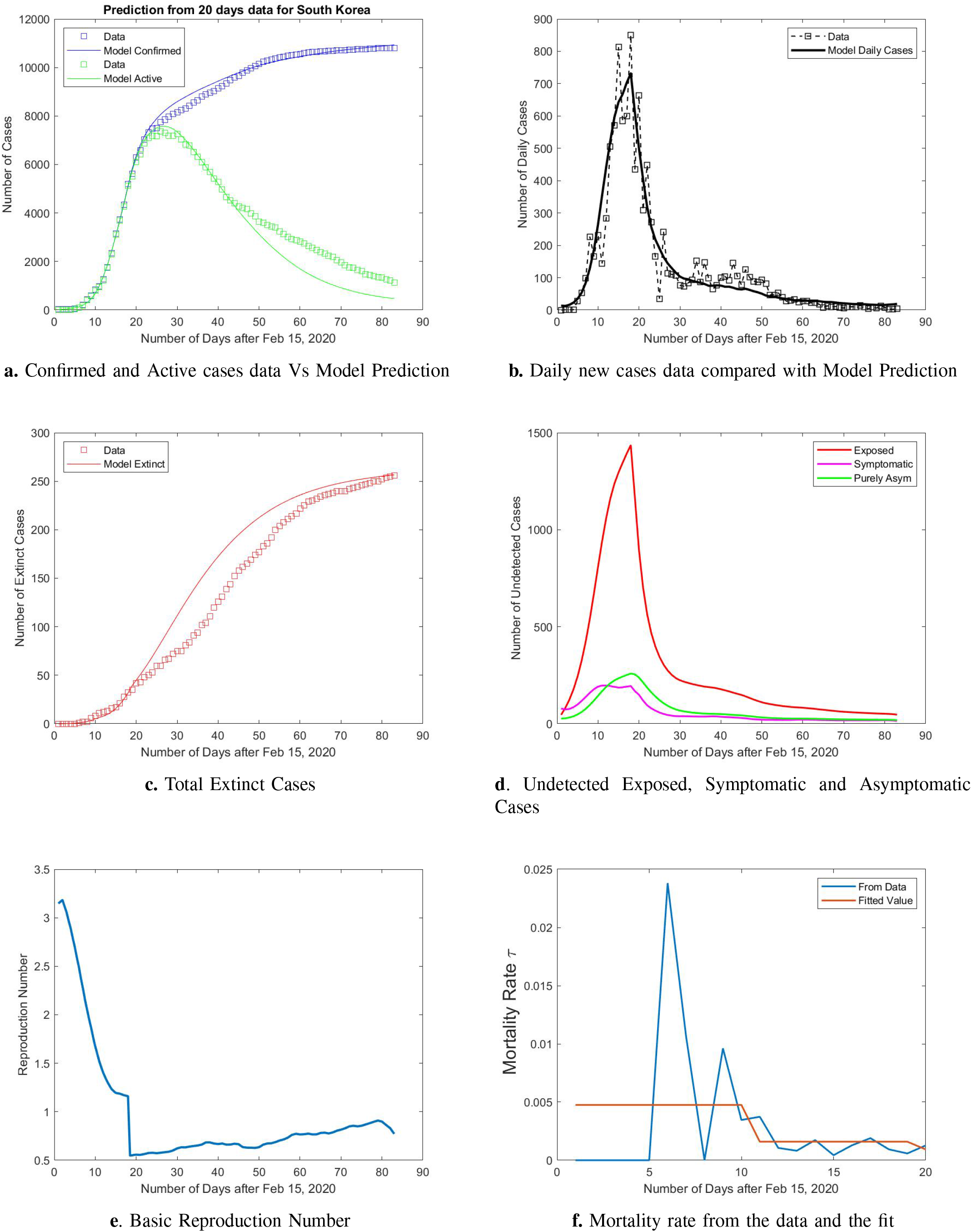
Model Predictions using first 20 days of data and comparison with real data for South Korea.

**Fig. 6:**
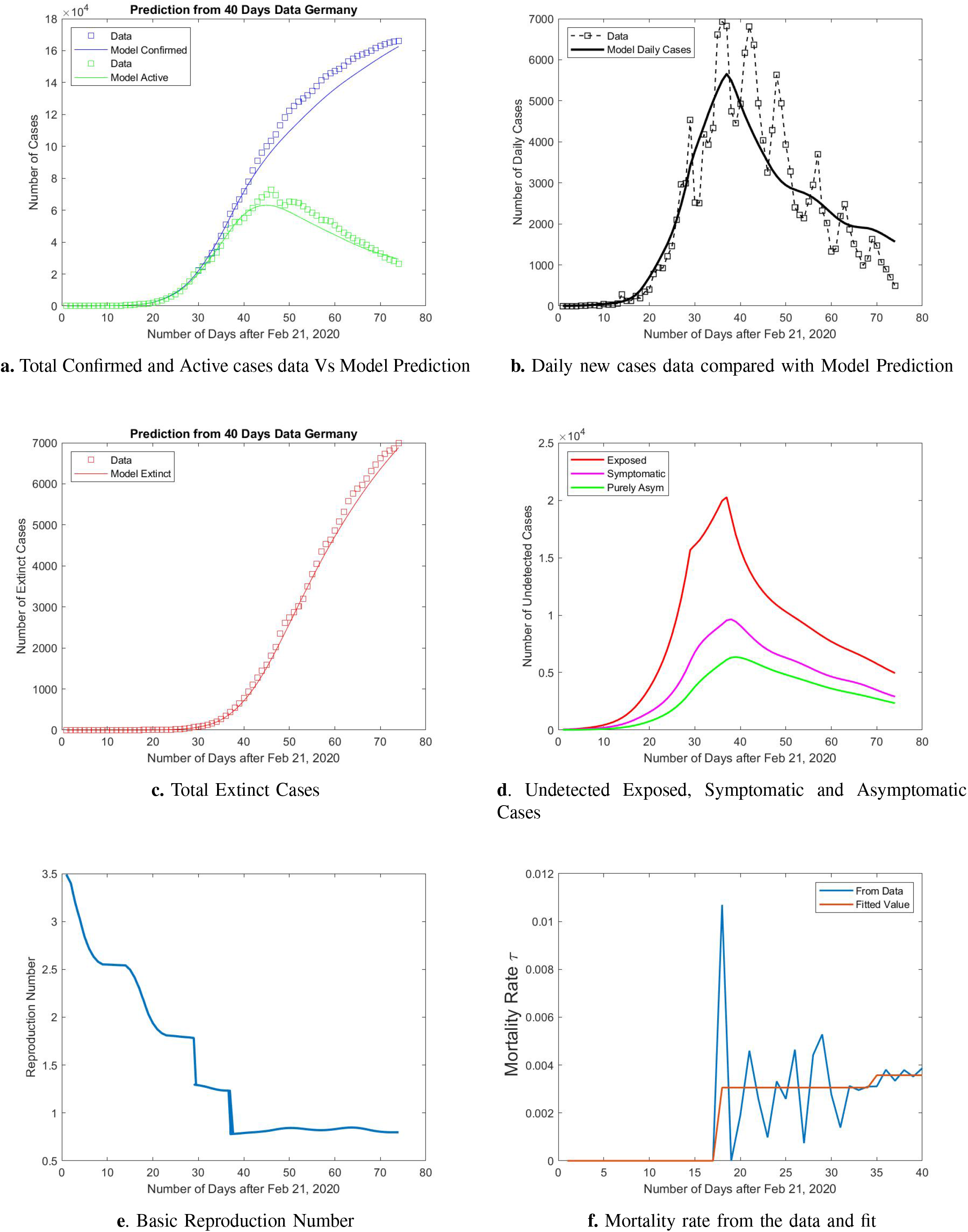
Model Predictions using first 40 days of data and comparison with real data for Germany.

### D. Effect of Increase in Testing

The factor by which an increase in testing can contain the infection is estimated for a relaxed lockdown situation. If there is 20% increase in the rate of transmission of infection than the current value due to this relaxation, then how fast the disease can be contained for 20k increase in tests per day is also plotted in Fig.4a. The daily new positive cases data and the prediction for the 10k increase in tests per day are plotted in Fig. 4c.

We also report a couple of additional scenarios for the prediction. If the lockdown is relaxed only for the month of June and due to that, if there is a 20% increase in the rate of transmission of infection than the current value, then how fast the disease will spread for 10k increase in tests per day is also plotted in Appendix Fig.9. If the lockdown is made stricter only for one month and if it is relaxed after one month, the evolution can be seen in Appendix Fig.10. The reproduction number is seen to go beyond one in the month of June for the relaxed lockdown. The infection to fatality ratio (IFR) is difficult to estimate during the course of the disease spread as the entities in the model are dynamically changing with time. We make a rough estimate by assuming that there is an average delay of 14 days between a person getting infected and becoming extinct.

The initial basic reproduction number for South Korea and Germany turns out to be 3.18 and 3.5, respectively, and for the USA it is 4.8. The South Korea reproduction number by our study is very close to 3.2 reported in [30] and the USA reproduction number is reported 4.2 on 16th March [3]. The USA basic reproduction number appears higher than the mean reported value [31] [32]. However, the Infection to Fatality Ratio (IFR) calculated with this high initial rate of transmission turns out to be around 0.7%, which is close to the reported value in [25].

A sensitivity study is carried out for the different parameters, as seen in Appendix Fig.11 and 12. The parameters are increased and decreased by 10% from the optimized values to see the changes in the outcomes.

## IV. Conclusion

Our findings show that reported cases in the USA could only be 16.5% of the total infected by June 1. If the lockdown is relaxed after June 15, it will lead to around 43k increase in total deaths, and doubling the everyday increase in testing from 10k to 20k can reduce this number by 13k. The model prediction shows that in the absence of a vaccine, the infection can last long till the end of this year and will infect around 5.35% of the total population, and the number of deaths could be around 166k if lockdown and social distancing conditions remain the same. Our simulation study predicts the future evolution of COVID-19 in the USA for various possible control measures in the coming months, including social distancing conditions and the number of tests per day, and thereby provides helpful inputs for policymakers.

## Data Availability

We collected the data from the following publicly available data sources: The total number of cases, active cases, daily new cases, and total and daily new deaths is collected from the worldometer[1]. Test per day data is collected from [27].Hospitalization data is collected from [21][24] .

https://www.worldometers.info/coronavirus

https://ourworldindata.org/grapher/full-list-covid-19-tests-per-day

https://gis.cdc.gov/grasp/covidnet/COVID19_3.html

https://covid19.healthdata.org/united-states-of-america

## Acknowledgement

AM would like to thank Dr. Shrikant Ambalkar, M.D for helpful discussions.

### APPENDIX

**Fig. 7:**
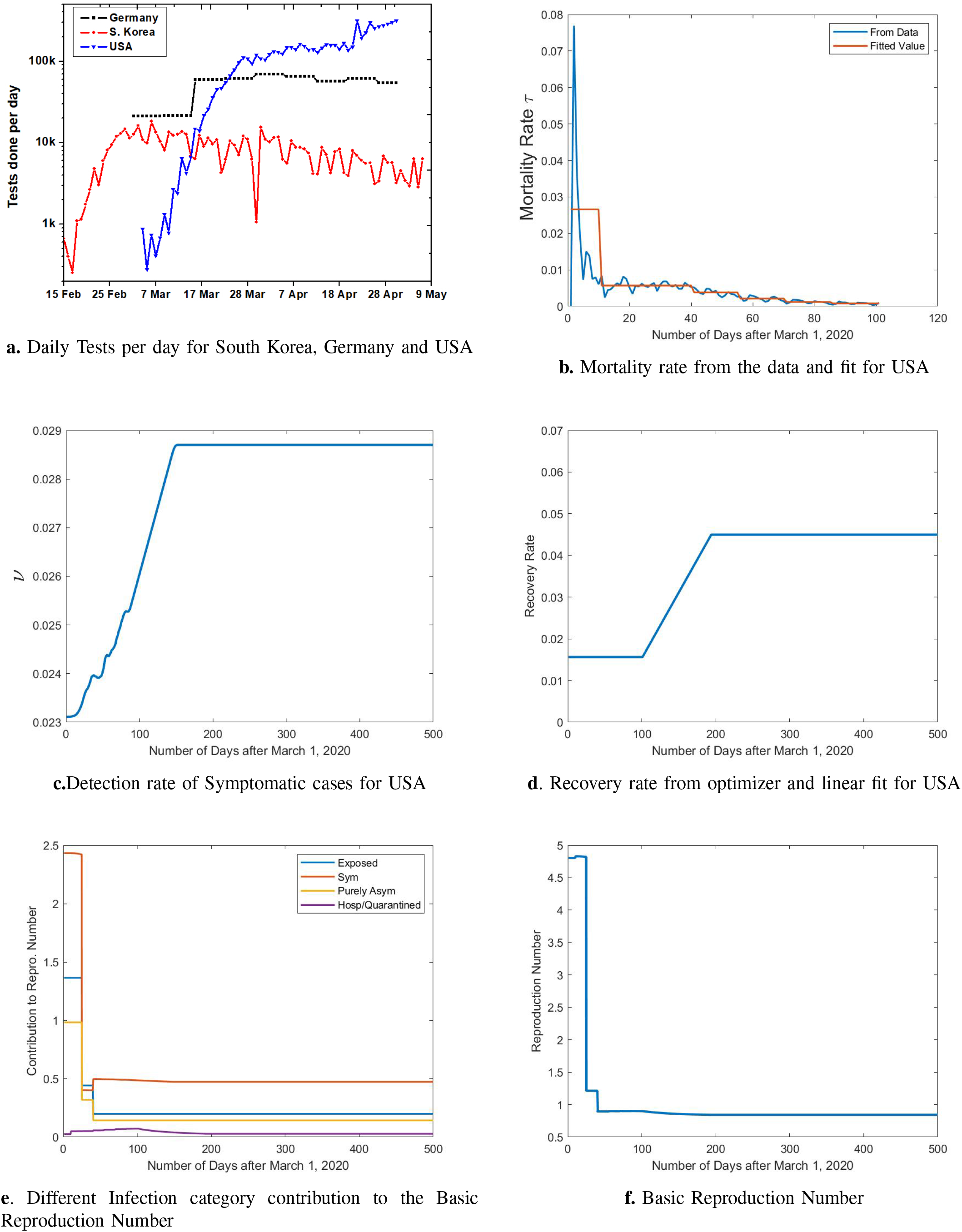
Tests per day, Time dependence of Model parameters and Reproduction Number for USA

**Fig. 8:**
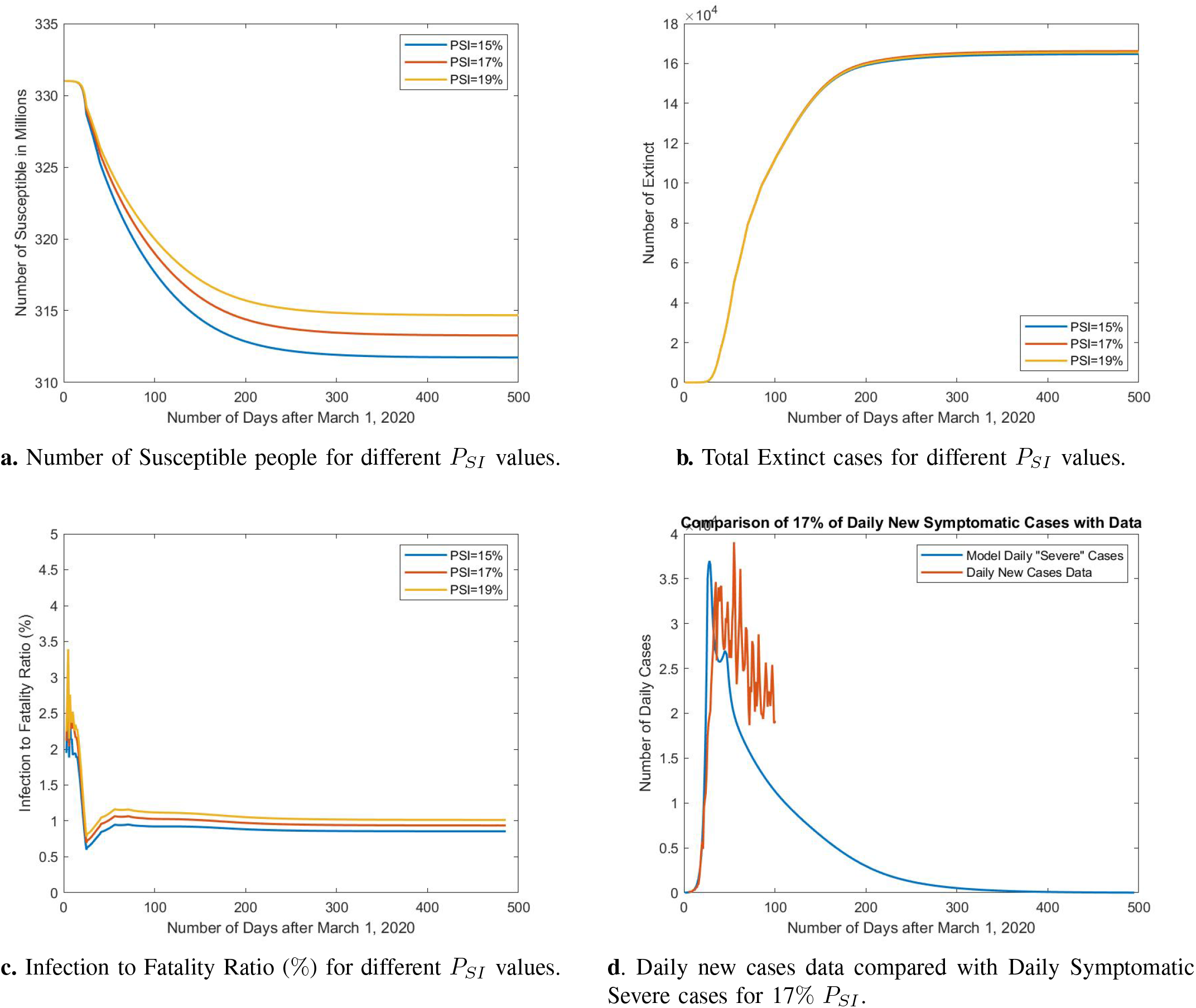
Sensitivity Study for the probability of Daily new Symptomatic Cases turning “Severe” for the USA

**Fig. 9:**
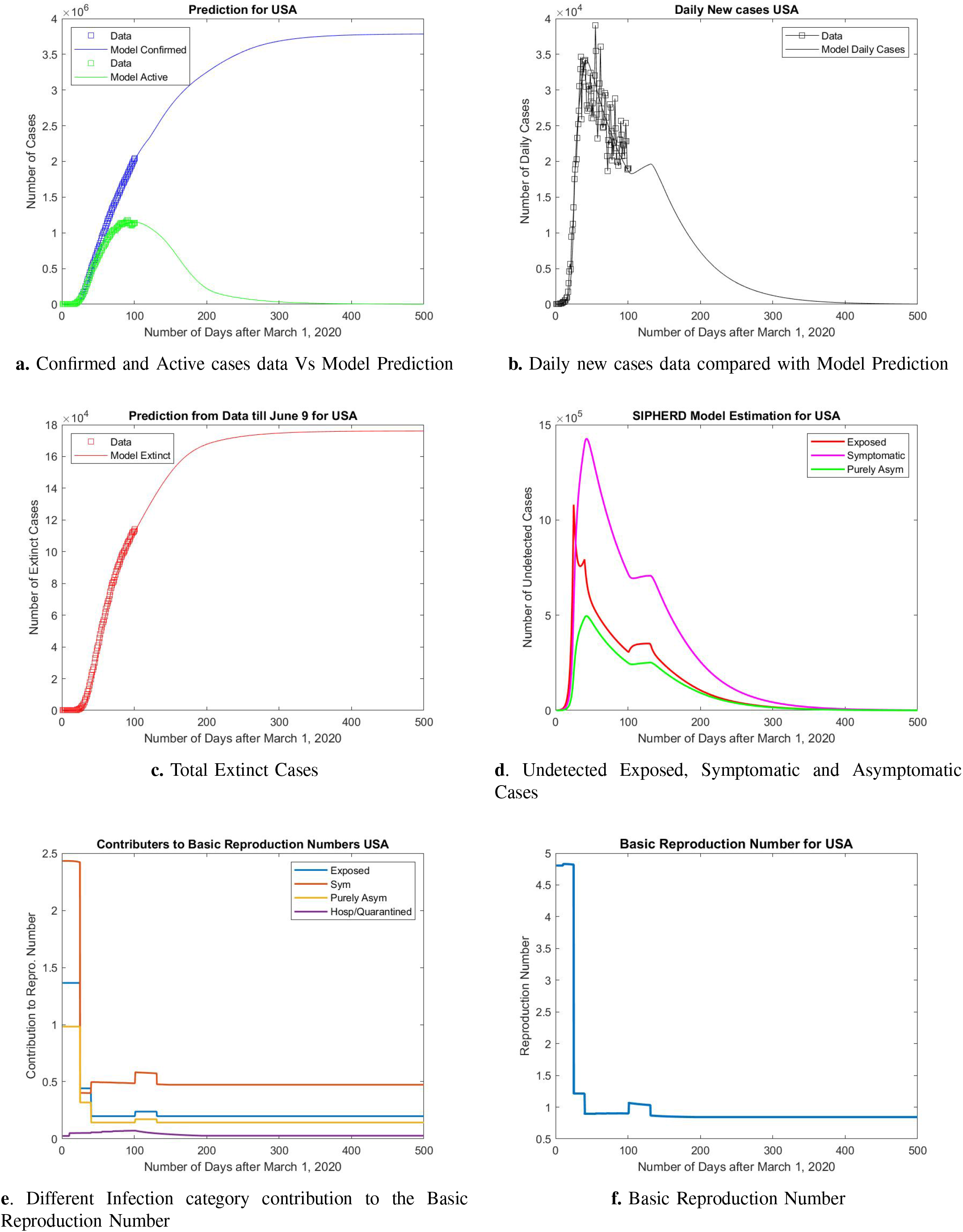
Model Predictions for USA if the lockdown conditions are relaxed after June 10 for a month.

**Fig. 10:**
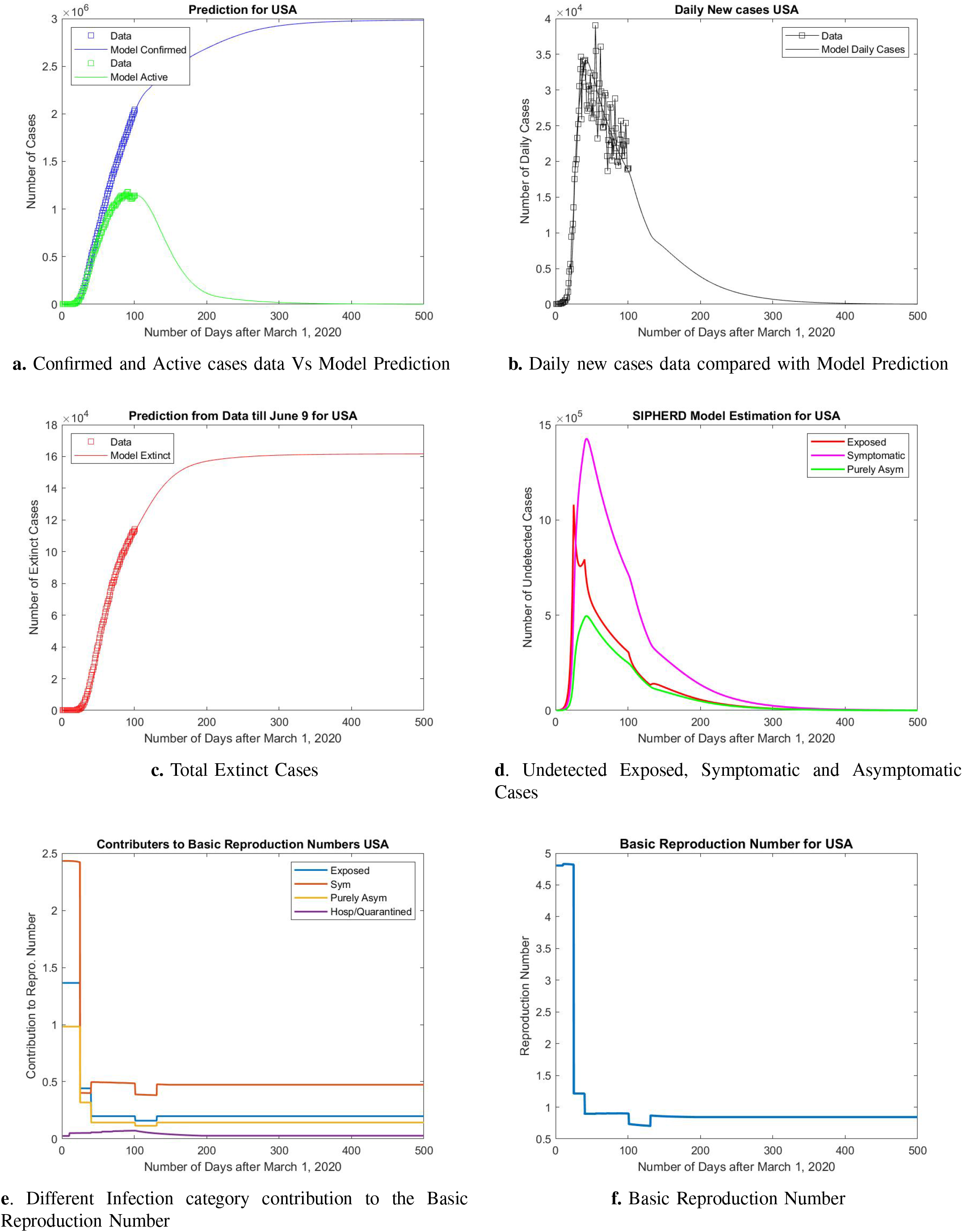
Model Predictions if the lockdown conditions are made more strict after June 10 for a month.

**Fig. 11:**
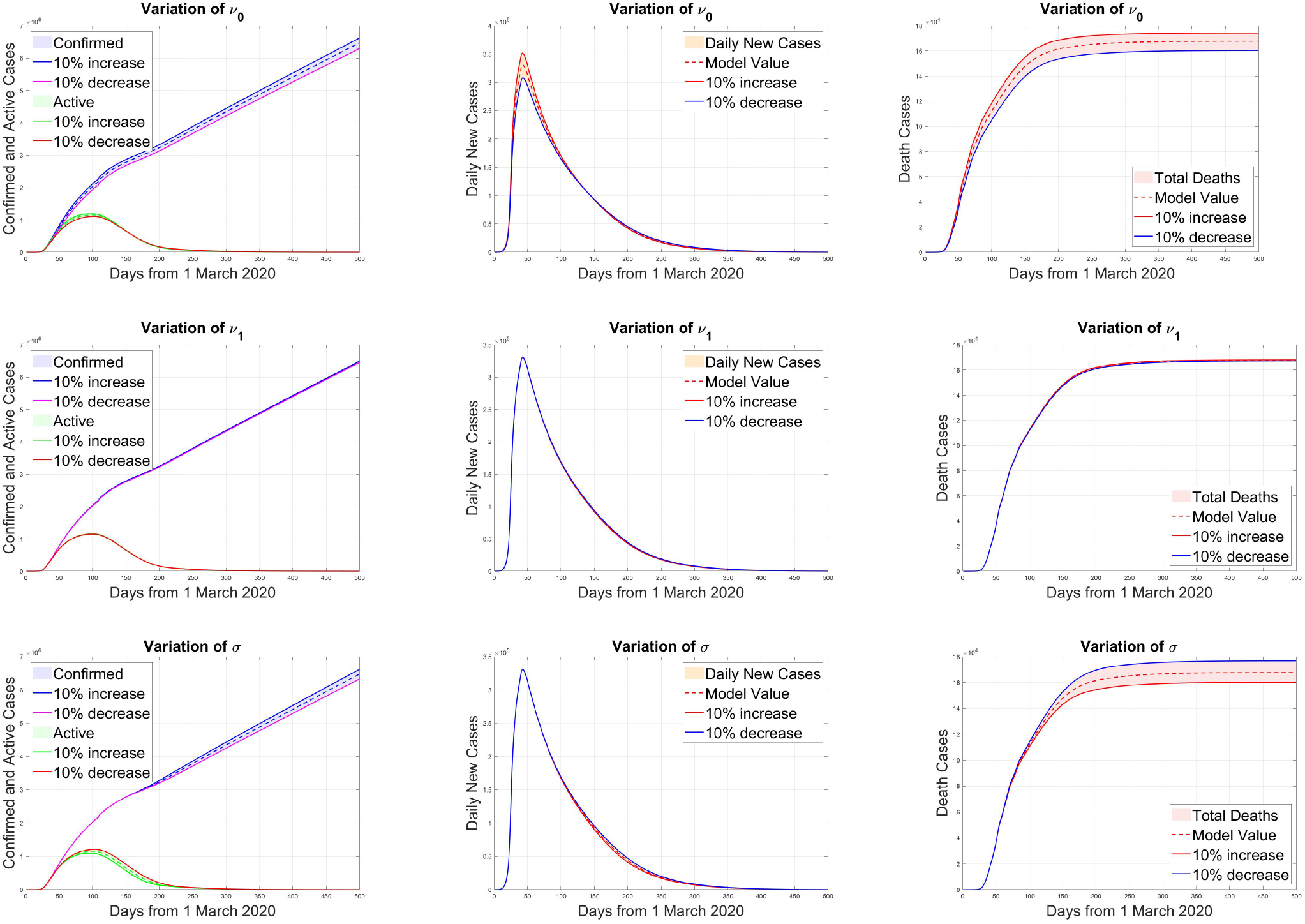
Sensitivity Study for the detection rate constants *ν*_0_, *ν*_1_ and recovery rate *s*. The Variation in total confirmed cases, total active cases and Extinct cases is plotted for 10% increase and decrease in the parameter value from the optimized value.

**Fig. 12:**
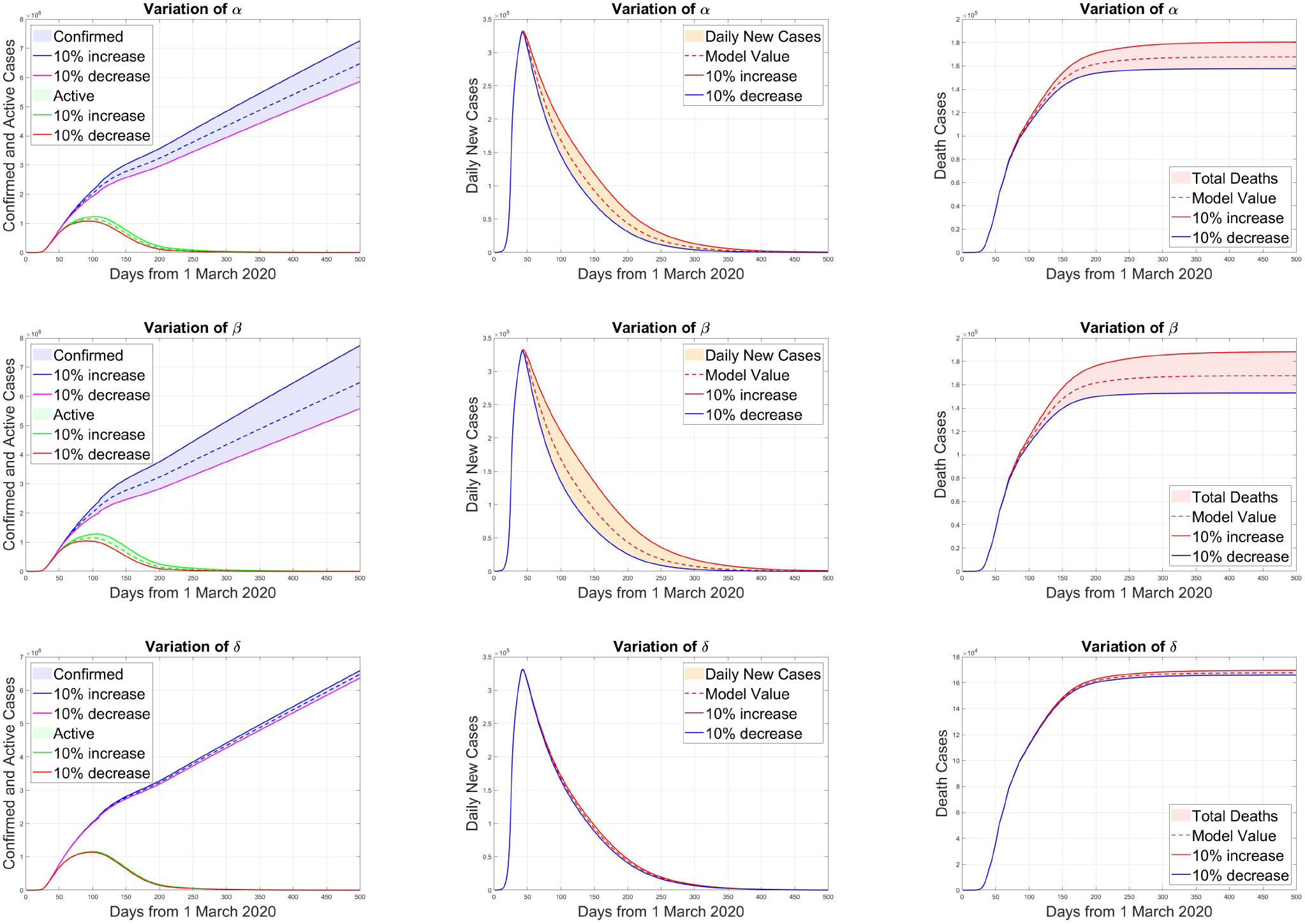
Sensitivity Study for the rate of transmission of infection *α,β* and *δ*. The Variation in total confirmed cases, total active cases and Extinct cases is plotted for 10% increase and decrease in the parameter value from the optimized value.

## Notes

### Competing Interest Statement

The authors have declared no competing interest.

### Funding Statement

This work is not supported by any funding agency.

### Author Declarations

This work does not need any approval of the IRB/oversight body.

## References

[1] https://www.worldometers.info/coronavirus.

[2] M. L. Holshue, C. DeBolt, S. Lindquist, K. H. Lofy, J. Wiesman, H. Bruce, C. Spitters, K. Ericson, S. Wilkerson, A. Tural et al., “First case of 2019 novel coronavirus in the united states,” New England Journal of Medicine, 2020.

[3] D. Gunzler and A. R. Sehgal, “Time-varying covid-19 reproduction number in the united states,” medRxiv, 2020.

[4] R. M. Anderson and R. M. May, Infectious diseases of humans: dynamics and control. Oxford university press, 1992.

[5] F. Brauer, C. Castillo-Chavez, and C. Castillo-Chavez, Mathematical models in population biology and epidemiology. Springer, 2012, vol. 2.

[6] H. W. Hethcote, “The mathematics of infectious diseases,” SIAM review, vol. 42, no. 4, pp. 599–653, 2000.

[7] O. Diekmann and J. A. P. Heesterbeek, Mathematical epidemiology of infectious diseases: model building, analysis and interpretation. John Wiley & Sons, 2000, vol. 5.

[8] S. M. Moghadas, A. Shoukat, M. C. Fitzpatrick, C. R. Wells, P. Sah, A. Pandey, J. D. Sachs, Z. Wang, L. A. Meyers, B. H. Singer et al., “Projecting hospital utilization during the covid-19 outbreaks in the united states,” Proceedings of the National Academy of Sciences, vol. 117, no. 16, pp. 9122–9126, 2020.

[9] J. J. Cavallo, D. A. Donoho, and H. P. Forman, “Hospital capacity and operations in the coronavirus disease 2019 (covid-19) pandemic—planning for the nth patient,” in JAMA Health Forum, vol. 1, no. 3. American Medical Association, 2020, pp. e200 345–e200 345.

[10] C. Wenham, “Modelling can only tell us so much: politics explains the rest,” The Lancet, vol. 395, no. 10233, p. 1335, 2020.

[11] K. Mizumoto, K. Kagaya, A. Zarebski, and G. Chowell, “Estimating the asymptomatic proportion of coronavirus disease 2019 (covid-19) cases on board the diamond princess cruise ship, yokohama, japan, 2020,” Eurosurveillance, vol. 25, no. 10, p. 2000180, 2020.

[12] C. Rothe, M. Schunk, P. Sothmann, G. Bretzel, G. Froeschl, C. Wallrauch, T. Zimmer, V. Thiel, C. Janke, W. Guggemos et al., “Transmission of 2019-ncov infection from an asymptomatic contact in germany,” New England Journal of Medicine, vol. 382, no. 10, pp. 970–971, 2020.

[13] B. Ivorra and A. M. Ramos, “Application of the be-codis mathematical model to forecast the international spread of the 2019–20 wuhan coronavirus outbreak,” ResearchGate http://dx.doi.org/10.13140/RG.2.2, vol. 31460, 2020.

[14] B. Ivorra, M. Ferrández, M. Vela-Pérez, and A. Ramos, “Mathematical modeling of the spread of the coronavirus disease 2019 (covid-19) considering its particular characteristics. the case of china,” Technical report, MOMAT, 03 2020, Tech. Rep., 2020.

[15] G. Giordano, F. Blanchini, R. Bruno, P. Colaneri, A. Di Filippo, A. Di Matteo, M. Colaneri et al., “A sidarthe model of covid-19 epidemic in italy,” arXiv preprint 2003.09861, 2020.

[16] D. Efimov and R. Ushirobira, “On an interval prediction of covid-19 development based on a seir epidemic model,” 2020.

[17] Y.-C. Chen, P.-E. Lu, C.-S. Chang, and T. Liu, “A time-dependent sir model for covid-19 with undetectable infected persons,” arXiv preprint 2003.00122, 2020.

[18] I. Covid, C. J. Murray et al., “Forecasting the impact of the first wave of the covid-19 pandemic on hospital demand and deaths for the usa and european economic area countries,” medRxiv, 2020.

[19] A. Mahajan, R. Solanki, and A. S. Namitha, “An epidemic model sipherd and its application for prediction of the spread of covid-19 infection for india and usa,” 2020.

[20] J. Zhang, M. Litvinova, W. Wang, Y. Wang, X. Deng, X. Chen, M. Li, W. Zheng, L. Yi, X. Chen et al., “Evolving epidemiology and transmission dynamics of coronavirus disease 2019 outside hubei province, china: a descriptive and modelling study,” The Lancet Infectious Diseases, 2020.

[21] C. Covid and R. Team, “Severe outcomes among patients with coronavirus disease 2019 (covid-19)—united states, february 12–march 16, 2020,” MMWR Morb Mortal Wkly Rep, vol. 69, no. 12, pp. 343–346, 2020.

[22] [Online]. Available: https://gis.cdc.gov/grasp/covidnet/COVID19_3.html

[23] S. Garg, “Hospitalization rates and characteristics of patients hospitalized with laboratory-confirmed coronavirus disease 2019—covid-net, 14 states, march 1–30, 2020,” MMWR. Morbidity and mortality weekly report, vol. 69, 2020.

[24] https://covid19.healthdata.org/united-states-of-america.

[25] H. Salje, C. T. Kiem, N. Lefrancq, N. Courtejoie, P. Bosetti, J. Paireau, A. Andronico, N. Hoze, J. Richet, C.-L. Dubost et al., “Estimating the burden of sars-cov-2 in france,” Science, 2020.

[26] C. P. E. R. E. Novel et al., “The epidemiological characteristics of an outbreak of 2019 novel coronavirus diseases (covid-19) in china,” Zhonghua liu xing bing xue za zhi= Zhonghua liuxingbingxue zazhi, vol. 41, no. 2, p. 145, 2020.

[27] https://ourworldindata.org/grapher/full-list-covid-19-tests-per-day.

[28] J. Wise, “Covid-19: Surveys indicate low infection level in community,” 2020.

[29] [Online]. Available: https://www.usnews.com/news/health-news/articles/2020-04-02/why-are-us-coronavirus-recovery-numbers-so-low

[30] Z. Zhuang, S. Zhao, Q. Lin, P. Cao, Y. Lou, L. Yang, S. Yang, D. He, and L. Xiao, “Preliminary estimating the reproduction number of the coronavirus disease (covid-19) outbreak in republic of korea and italy by 5 march 2020,” International Journal of Infectious Diseases, 2020.

[31] Y. Liu, A. A. Gayle, A. Wilder-Smith, and J. Rocklöv, “The reproductive number of covid-19 is higher compared to sars coronavirus,” Journal of travel medicine, 2020.

[32] J. M. Read, J. R. Bridgen, D. A. Cummings, A. Ho, and C. P. Jewell, “Novel coronavirus 2019-ncov: early estimation of epidemiological parameters and epidemic predictions,” MedRxiv, 2020.

